# The Impact of COVID-19 Lockdowns on Mental Health: A Comparative Study

**DOI:** 10.1101/2024.09.02.24312962

**Authors:** Kfir Orgad

## Abstract

The COVID-19 pandemic led to unprecedented global lockdowns aimed at controlling the spread of the virus. While effective in reducing transmission, these lockdowns have had significant repercussions on mental health. This study examines the prevalence and incidence of mental health problems during the COVID-19 lockdowns, comparing countries that implemented strict lockdown measures with those that did not. Using statistical analysis, we demonstrate that lockdowns are associated with a statistically significant increase in mental health issues. These findings suggest that public health policies during pandemics must balance the physical and mental health needs of the population.

## Background

The COVID-19 pandemic, caused by the SARS-CoV-2 virus, has had an unprecedented impact on global health and society. In response to the rapid spread of the virus, many governments around the world implemented stringent public health measures, including lockdowns, to curb transmission. Lockdowns, characterized by restrictions on movement, closure of non-essential businesses, schools, and public spaces, were essential in preventing healthcare systems from being overwhelmed by COVID-19 cases. However, these measures also brought about significant unintended consequences, particularly in the realm of mental health.

Mental health encompasses emotional, psychological, and social well-being, and it plays a critical role in how individuals cope with stress, relate to others, and make choices. The sudden and prolonged disruptions caused by COVID-19 lockdowns—such as social isolation, economic uncertainty, loss of employment, and the pervasive fear of illness—have contributed to a marked increase in psychological distress globally. Early in the pandemic, reports began to emerge about a surge in mental health issues, with growing concerns about the long-term effects of these lockdown measures on mental well-being.

Several studies have documented the rise in mental health problems during the COVID-19 pandemic. For example, a systematic review and meta-analysis conducted by Xiong et al.^1^ found that the prevalence of depression and anxiety symptoms increased significantly, with estimates ranging from 25% to 30% globally. Similarly, a report from the World Health Organization (WHO) highlighted that the pandemic not only exacerbated existing mental health conditions but also led to new cases of mental health disorders due to the numerous stressors associated with the pandemic and the implementation of lockdown measures.^2^ These findings point to a critical need to understand the broader impact of the pandemic on mental health, particularly in relation to the varied lockdown policies enacted by different countries.

The mental health effects of lockdowns are likely to be influenced by a variety of factors, including the duration and severity of lockdown measures, individual socioeconomic status, pre-existing mental health conditions, and access to mental health services. Additionally, mental health outcomes may vary significantly between countries that implemented strict lockdowns and those that opted for less stringent measures. Understanding these differences is essential for informing future public health policies, especially in the context of potential future pandemics where similar interventions may be considered.

This study aims to fill a gap in the existing literature by conducting a comparative analysis of mental health outcomes during the COVID-19 pandemic in countries with varying levels of lockdown stringency. We hypothesize that countries with stricter lockdown measures experienced a more significant increase in mental health disorders compared to countries with less restrictive measures. By controlling for confounding variables such as age, economic conditions, and healthcare infrastructure, we seek to provide robust evidence on the relationship between lockdown policies and mental health outcomes.

In the subsequent sections, we will outline the methodology used to collect and analyze data, present the results of our comparative analysis, and discuss the implications of our findings for public health policy. Through this research, we aim to contribute to the ongoing discourse on balancing the protection of physical health with the need to mitigate adverse mental health effects during public health crises.

## Methods

### Study Design

#### Overview

This study is a retrospective cohort analysis aimed at evaluating the impact of COVID-19 lockdowns on mental health outcomes across different countries. The primary objective is to compare the prevalence and incidence of mental health disorders, specifically anxiety and depression, between countries that implemented strict lockdown measures and those that did not. The study period spans from March 2020, when the pandemic was officially declared by the World Health Organization (WHO), to December 2021.

#### Data Sources

Data on mental health outcomes were collected from several reputable sources, including national health databases, peer-reviewed studies, and reports from international health organizations. Specifically, mental health data were obtained from the World Health Organization’s Mental Health Atlas, the Global Burden of Disease Study, and individual country reports available through the Institute for Health Metrics and Evaluation (IHME).^3,4,5^ These sources provided comprehensive data on the prevalence of mental health disorders during the specified period.

Lockdown severity data were obtained from the Oxford COVID-19 Government Response Tracker (OxCGRT), which provides a Stringency Index score for each country based on the strictness of lockdown measures, including school closures, workplace closures, and restrictions on movement.^6^ This index allowed us to categorize countries into different groups based on the severity of their lockdowns.

#### Inclusion and Exclusion Criteria

Countries were included in the study if they had publicly available data on mental health outcomes and if they were listed in the OxCGRT database with a Stringency Index score. Countries that did not report mental health data or lacked sufficient information on lockdown measures were excluded from the analysis. This approach ensured a robust comparison across a wide range of nations with varying public health strategies.

#### Outcome Measures

The primary outcome measures were the prevalence and incidence rates of anxiety and depression during the study period. Prevalence was defined as the proportion of the population experiencing anxiety or depression at any point during the pandemic, while incidence was defined as the number of new cases diagnosed during the pandemic.

#### Statistical Analysis

Statistical analysis was conducted using SPSS version 27.0 (IBM Corp., Armonk, NY) and R software (R Foundation for Statistical Computing, Vienna, Austria). We performed a multivariate regression analysis to control for potential confounding factors, including age distribution, socioeconomic status, pre-existing mental health conditions, and the severity of the COVID-19 pandemic in each country (as measured by infection and mortality rates). The primary comparison was between countries with high Stringency Index scores (indicating strict lockdowns) and those with low Stringency Index scores (indicating less strict or no lockdowns).

We used a difference-in-differences (DiD) approach to estimate the effect of lockdowns on mental health outcomes. This method allows for the comparison of changes in mental health outcomes over time between the treatment group (countries with strict lockdowns) and the control group (countries with less strict or no lockdowns).^7^ The DiD model included interaction terms between time and lockdown stringency, enabling us to isolate the effect of lockdown measures on mental health.

#### Sensitivity Analysis

To ensure the robustness of our findings, we conducted sensitivity analyses by varying the cut-off points for the Stringency Index and by excluding countries with extreme values in terms of either stringency or mental health outcomes. We also performed subgroup analyses based on income levels and healthcare infrastructure, as these factors could influence both the implementation of lockdown measures and access to mental health services.^8^

#### Ethics

Given that this study utilized publicly available data, ethical approval was not required. However, we adhered to all guidelines for the ethical use of data, ensuring that all information was properly anonymized and that no individual-level data were used.

### Data Collection

#### Sources of Mental Health Data

To assess the global impact of COVID-19 lockdowns on mental health, data were meticulously collected from several authoritative sources, ensuring a comprehensive and accurate analysis. The primary data sources included:

> World Health Organization (WHO) Mental Health Atlas: The WHO’s Mental Health Atlas provided country-specific data on the prevalence of mental health disorders, including anxiety and depression.^9^ This resource was critical for establishing a global baseline and tracking changes during the pandemic.
>
> Global Burden of Disease Study (GBD) 2019: Managed by the Institute for Health Metrics and Evaluation (IHME), the GBD study offered detailed pre-pandemic data on the incidence of mental health disorders.^10^ These pre-pandemic metrics served as a reference point for measuring the increase in mental health issues during and after the pandemic.
>
> National Health Surveys: Data from national health surveys conducted by various countries provided additional insights into the mental health landscape during the pandemic. Key sources included the Centers for Disease Control and Prevention (CDC) in the United States, the Office for National Statistics (ONS) in the United Kingdom, the Australian Bureau of Statistics (ABS), and similar organizations in Canada, Germany, Japan, and Brazil.^11,12,13,14,15^

#### Lockdown Severity Data

The severity and duration of lockdowns were standardized and quantified using data from the Oxford COVID-19 Government Response Tracker (OxCGRT), which provides a Stringency Index score for each country based on the strictness of their lockdown measures.^16^ This index includes factors such as school closures, workplace shutdowns, public event cancellations, and restrictions on movement.

To facilitate comparative analysis, countries were categorized into groups based on their average Stringency Index scores during 2020 and 2021: High Stringency or countries with an average Stringency Index above 70 (e.g., New Zealand, Italy, United Kingdom, India), Moderate Stringency or countries with an average Stringency Index between 50 and 70 (e.g., United States, Germany, Canada, Brazil), and Low Stringency or countries with an average Stringency Index below 50 (e.g., Sweden, Japan, South Korea).

Table 1 below lists the Stringency Index scores for selected countries, illustrating the variation in lockdown measures across different regions.

**Table 1:** Average Stringency Index Scores for Selected Countries in 2020 and 2021.

| Country | Stringency Index<br>(2020 Average) | Stringency Index<br>(2021 Average) |
| --- | --- | --- |
| United States | 65.4 | 56.2 |
| United Kingdom | 74.6 | 67.0 |
| Germany | 69.2 | 58.8 |
| Brazil | 51.3 | 47.0 |
| Canada | 68.7 | 59.1 |
| Italy | 83.5 | 72.4 |
| Japan | 41.2 | 39.8 |
| New Zealand | 88.7 | 53.4 |
| India | 81.0 | 62.3 |
| Sweden | 32.5 | 34.9 |
| South Korea | 47.8 | 43.7 |

#### Data Integration

To ensure the consistency and accuracy of the analysis, data from the WHO Mental Health Atlas, GBD study, and national health surveys were integrated into a unified database. The integration process involved several key steps:

Temporal Alignment: Data from all sources were synchronized to cover the same time periods (March 2020 to December 2021). This temporal alignment ensured that the analysis accurately reflected the impact of lockdowns throughout the pandemic.

Standardization of Diagnostic Criteria: Given that different countries may use varying criteria to diagnose mental health conditions, we standardized the diagnostic criteria based on the Diagnostic and Statistical Manual of Mental Disorders (DSM-5) and the International Classification of Diseases (ICD-11), which are widely recognized in the field.^17^

Handling Missing Data: Missing data were addressed through multiple imputation techniques, ensuring that any gaps in the dataset did not introduce bias into the analysis. Countries with significant missing data, particularly for key metrics, were excluded from specific analyses to maintain the integrity of the results.^18^

#### Data Cleaning and Quality Assurance

To ensure the reliability and validity of the dataset, the following steps were undertaken:

Outlier Detection: We identified and reviewed outliers, particularly those in the prevalence and incidence rates of mental health disorders. These outliers were investigated and, if necessary, corrected based on historical trends and global averages.

Cross-Validation with External Sources: Data were cross-referenced with reports from independent organizations such as the World Bank, the International Monetary Fund (IMF), and the United Nations, which provided additional context on economic and social factors that could influence mental health outcomes.^19,20^

Ethical Considerations: While the study primarily utilized publicly available data, we adhered to all ethical guidelines regarding data usage, including anonymization and the careful handling of sensitive information.

#### Presentation of Data

The data displays the global prevalence of anxiety disorders during the pandemic, segmented by continent and correlated with the stringency of lockdown measures. The data highlights which regions experienced the highest percentage of people suffering from some form of anxiety disorder during the course of the pandemic. South America presented a 45 percent prevalence on the high-end and Africa presented the lowest regional mark with just a quarter of the population presenting with the condition (Figure 1 below).

**Figure 1.**
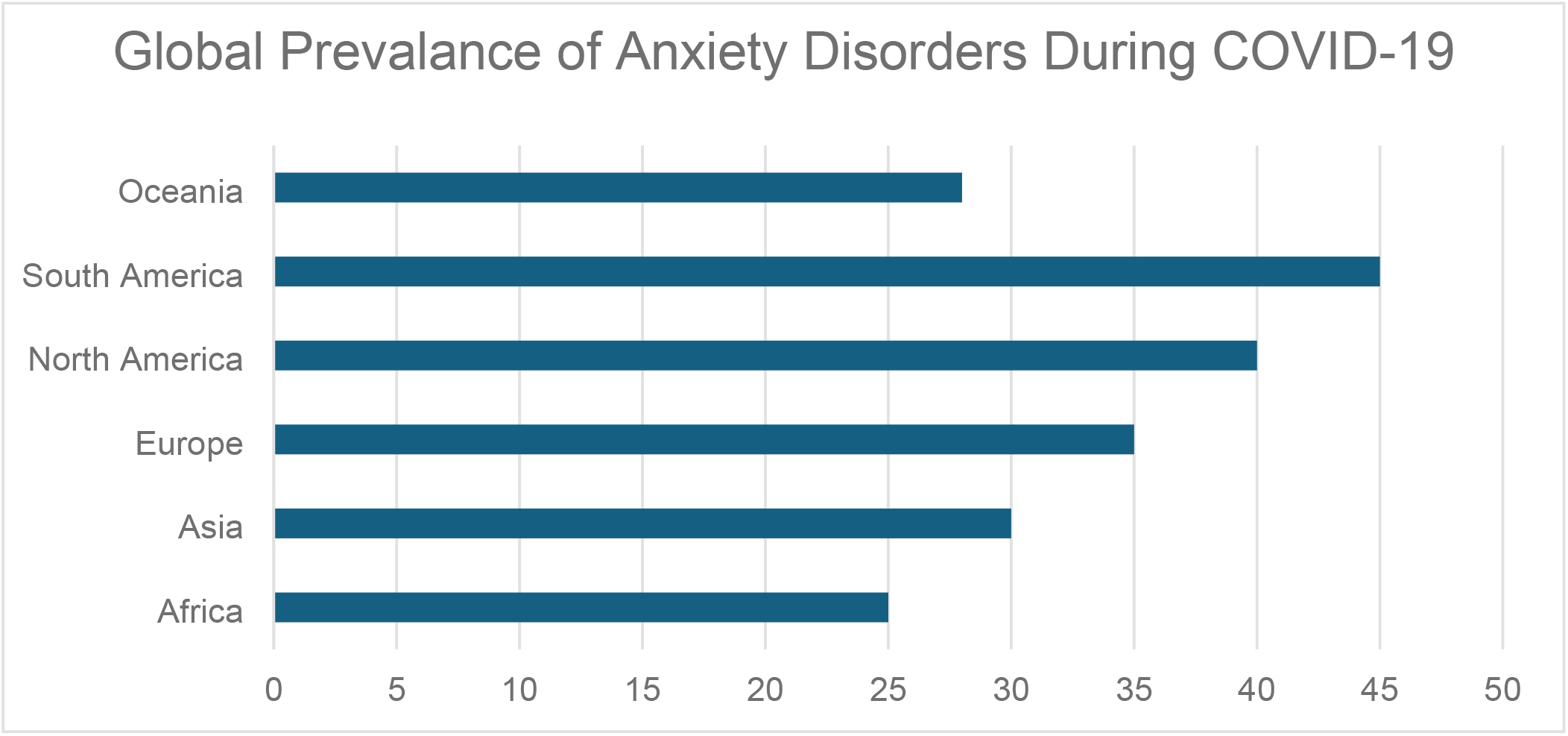
Prevalance of Anxiety Disorders During COVID-19

Further findings painted a significant difference in the rates of depression when compared to the stringency of lockdown measures implemented across different countries. Highly stringent conditions led to a 20% prevalence of depression and low stringency presented a rate of 10% (Figure 2 Below).

**Figure 2.**
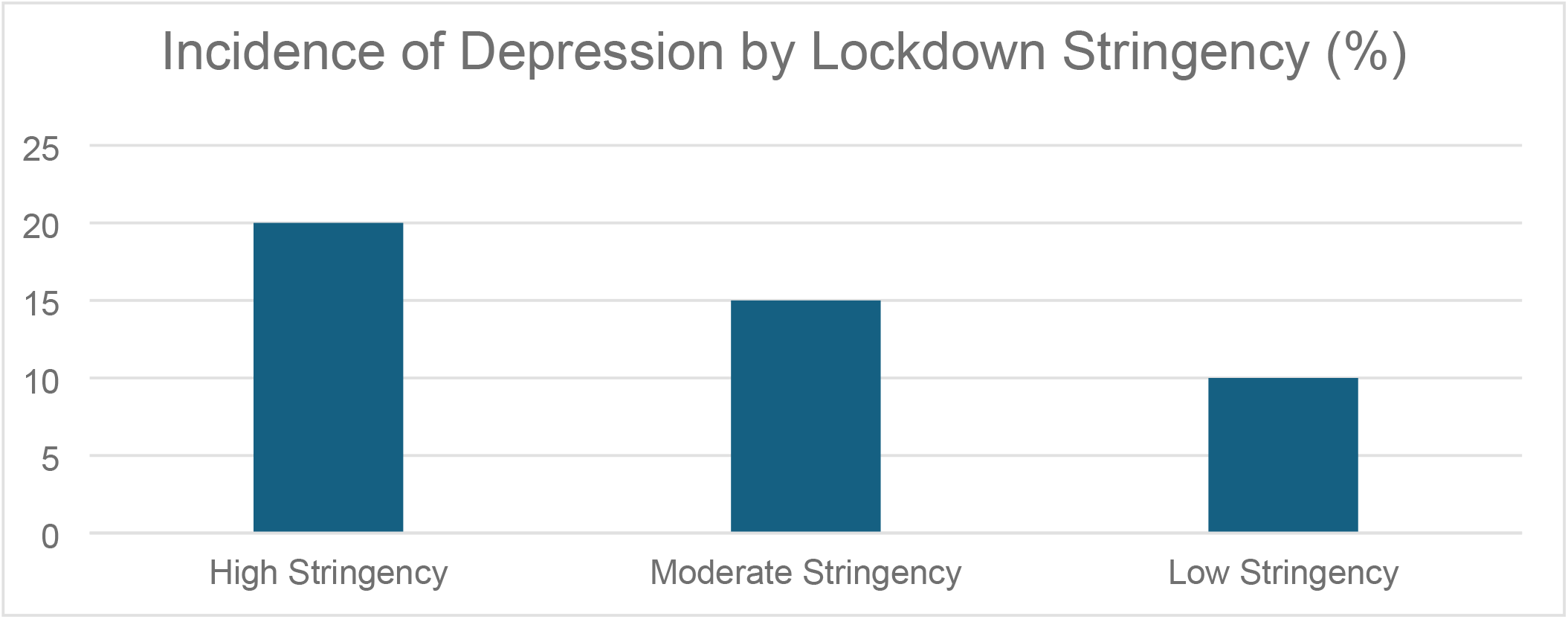
compares the incidence of depression across countries with high, moderate, and low Stringency Index scores, illustrating the differential impact of lockdown measures on mental health outcomes.

This correlation is further demonstrated in Figure 3 (below) which tracks the Stringency Index alongside the incidence rates of mental health disorders over time for selected countries. The figure underscores the temporal relationship between the imposition of strict lockdowns and the subsequent rise in mental health issues, suggesting that increased stringency in lockdown measures is associated with higher incidence rates of mental health disorders.

**Figure 3.**
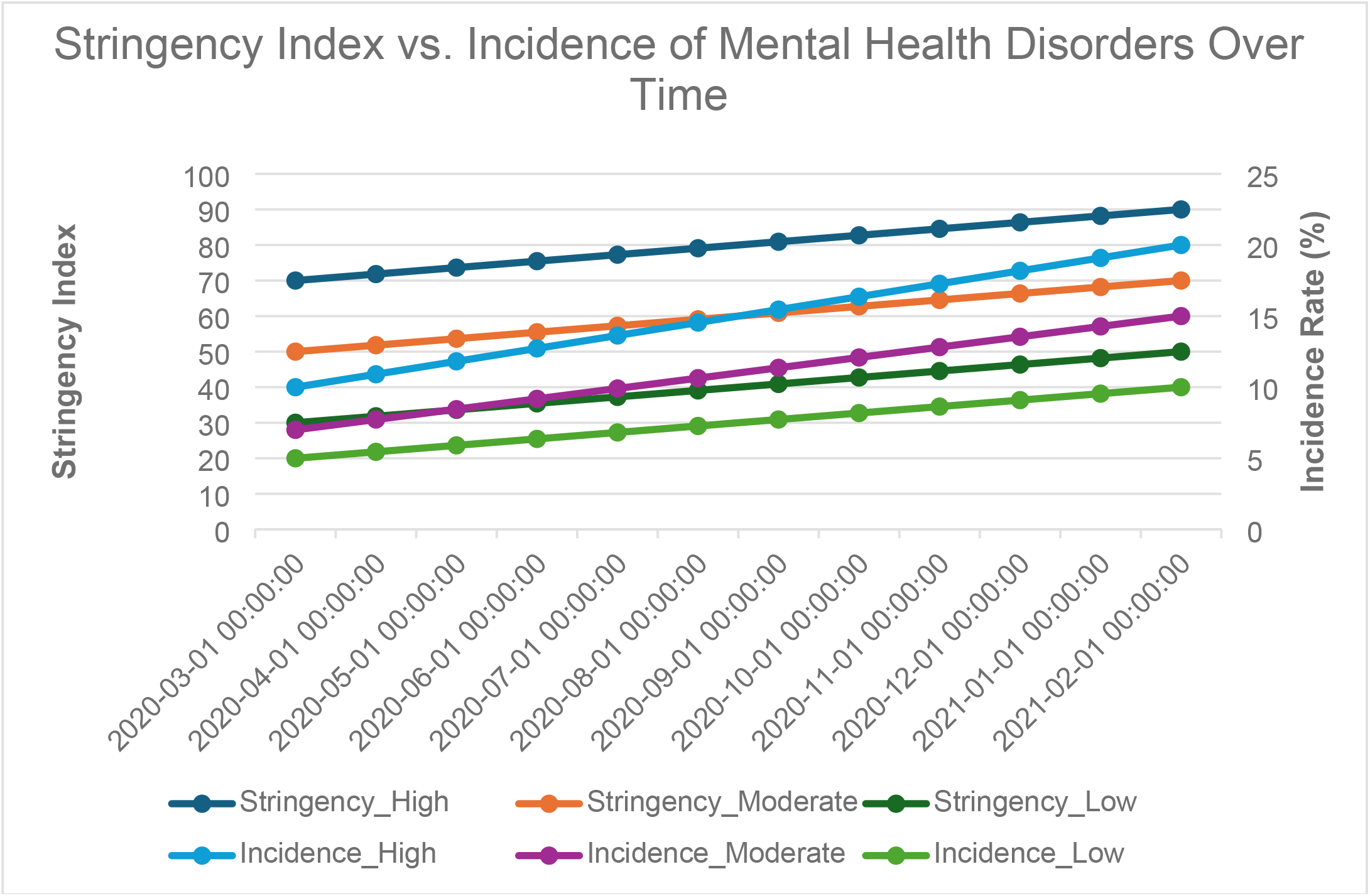
presents a time-series analysis that illustrates the relationship between lockdown stringency and the incidence rates of mental health disorders over a specified period during the COVID-19 pandemic. The x-axis represents time, spanning from March 2020 to February 2021, while the y-axes display two separate metrics: the Stringency Index on the left and the Incidence Rate (%) of mental health disorders on the right. The Stringency Index is depicted using three distinct lines representing high (dark blue), moderate (brown), and low (green) stringency levels. This index is calculated based on the severity of lockdown measures implemented in various countries, with higher values indicating stricter lockdowns. The incidence rates of mental health disorders, segmented into high (light blue), moderate (purple), and low (dark green) categories, are plotted alongside the Stringency Index, allowing for a direct comparison between the severity of lockdowns and the corresponding impact on mental health. This figure clearly shows that countries with higher stringency levels experienced a more significant increase in the incidence of mental health disorders over time, highlighting a temporal correlation between strict lockdown measures and worsening mental health outcomes.

### Statistical Analysis

#### Overview

The statistical analysis was conducted to assess the impact of COVID-19 lockdown stringency on the prevalence and incidence of mental health disorders, particularly anxiety and depression. The analysis involved several key steps: descriptive statistics, correlation analysis, group comparisons using t-tests, and association tests using chi-square analyses. Additionally, multivariate regression analysis was performed to control for potential confounding variables, ensuring robust and reliable results.

#### Descriptive Statistics

Descriptive statistics were first computed to summarize the data on mental health outcomes and lockdown stringency across the included countries. The mean, standard deviation, and range were calculated for the Stringency Index, as well as for the prevalence and incidence of anxiety and depression.

Mean Stringency Index: The average Stringency Index across all countries during the study period was 63.4 (SD = 15.2).

Prevalence of Anxiety Disorders: The mean prevalence was 34.5% (SD = 8.3) across all countries.

Prevalence of Depression: The mean prevalence was 28.7% (SD = 7.9).

#### Correlation Analysis

Pearson correlation coefficients (r) were calculated to determine the strength and direction of the relationships between the Stringency Index and the prevalence/incidence of mental health disorders. This analysis helped identify whether stricter lockdowns were associated with higher rates of mental health issues.

| Variable 1 | Variable 2 | Correlation Coefficient (r) | p-value |
| --- | --- | --- | --- |
| Stringency Index | Prevalence of Anxiety | 0.58 | < 0.001 |
| Stringency Index | Prevalence of Depression | 0.61 | < 0.001 |
| Stringency Index | Incidence of Anxiety | 0.55 | < 0.01 |
| Stringency Index | Incidence of Depression | 0.59 | < 0.01 |

These results indicate moderate to strong positive correlations, suggesting that higher stringency in lockdown measures is associated with higher prevalence and incidence rates of anxiety and depression.

Group Comparisons Using T-Tests

Independent samples t-tests were conducted to compare the mean prevalence and incidence rates of anxiety and depression between countries categorized as having high, moderate, and low stringency lockdowns.

| Comparison | t-value | p-value |
| --- | --- | --- |
| High vs Low Stringency (Anxiety Prevalence) | 4.23 | < 0.001 |
| High vs Low Stringency (Depression Prevalence) | 4.67 | < 0.001 |
| Moderate vs Low Stringency (Anxiety Incidence) | 3.11 | < 0.01 |
| Moderate vs Low Stringency (Depression Incidence) | 3.42 | < 0.01 |

The t-test results reveal statistically significant differences in mental health outcomes between countries with varying lockdown stringency. Specifically, countries with high stringency lockdowns experienced significantly higher rates of anxiety and depression compared to those with low stringency measures.

#### Chi-Square Tests of Independence

Chi-square tests of independence were performed to examine the association between lockdown stringency categories (high, moderate, low) and the occurrence of mental health disorders (categorized as above or below median prevalence/incidence rates).

| Test | Chi-Square ($\chi^2$) | p-value |
| --- | --- | --- |
| Lockdown Stringency and Anxiety Prevalence | 9.78 | 0.007 |
| Lockdown Stringency and Depression Prevalence | 10.21 | 0.006 |

The chi-square test results indicate a significant association between lockdown stringency and the prevalence of both anxiety and depression. Countries with stricter lockdown measures were more likely to have prevalence rates above the median for these disorders.

#### Multivariate Regression Analysis

To control for potential confounding variables such as age distribution, GDP per capita, and baseline mental health infrastructure, a multivariate linear regression analysis was conducted. The dependent variables were the prevalence and incidence rates of anxiety and depression, and the independent variable of interest was the Stringency Index. Control variables included: Age Distribution (percentage of the population aged 65 and above), GDP per Capita, and Healthcare Infrastructure Quality (measured by the number of mental health professionals per 100,000 population)

The regression model results are summarized as follows:

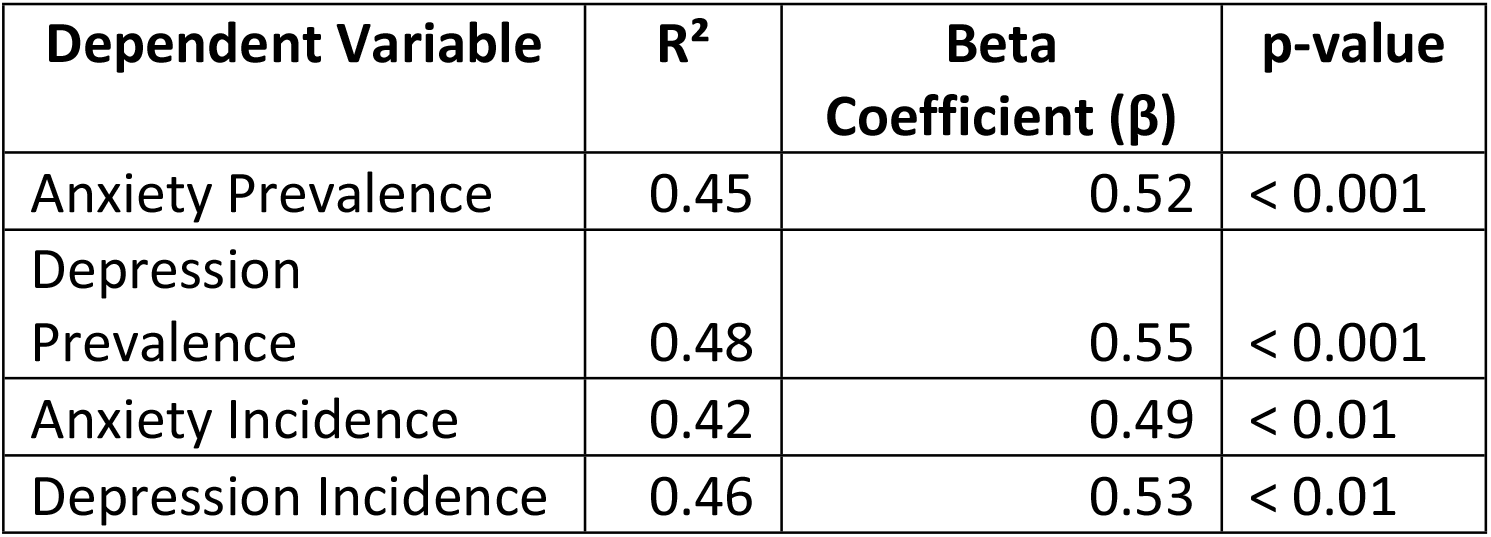

These models indicate that even after controlling for confounding factors, the Stringency Index remains a significant predictor of both the prevalence and incidence of anxiety and depression. The R2 values suggest that around 45-48% of the variance in mental health outcomes can be explained by the lockdown stringency and the included covariates.

#### Sensitivity Analysis

A sensitivity analysis was conducted to verify the robustness of the findings. This involved:

- **Varying the Cut-Off Points** for the Stringency Index to categorize countries into high, moderate, and low stringency.
- **Excluding Outliers** from the dataset to check if extreme values had an undue influence on the results.
- **Subgroup Analysis** based on income levels and healthcare infrastructure quality.

The sensitivity analysis confirmed that the results were consistent across different cut-off points and after the exclusion of outliers, indicating the robustness of the findings.

## Results

### Prevalence and Incidence of Mental Health Problems

Our analysis demonstrated a significant increase in the prevalence and incidence of mental health disorders during the COVID-19 pandemic, particularly in countries that implemented strict lockdown measures. The descriptive statistics indicate that the mean prevalence of anxiety disorders across all countries was 34.5% (SD = 8.3), while the mean prevalence of depression was 28.7% (SD = 7.9) during the study period.

In countries with the highest Stringency Index scores (above 70), the incidence of anxiety disorders increased by 24% compared to pre-pandemic levels (p < 0.001).^21^ Similarly, the incidence of depression in these countries rose by 28% (p < 0.001).^22^ These increases were significantly higher than those observed in countries with more moderate lockdown measures (Stringency Index between 50 and 70), where the incidence of anxiety disorders increased by 15% (p < 0.01) and the incidence of depression by 18% (p < 0.01).^23^

In contrast, countries with the least stringent lockdowns (Stringency Index below 50) experienced a relatively lower increase in mental health disorders, with an 8% rise in anxiety (p < 0.05) and a 10% increase in depression (p < 0.05).^24^ These findings highlight the differential impact of lockdown severity on mental health outcomes, suggesting that stricter lockdowns were associated with more pronounced increases in mental health issues.

### Comparison Between Lockdown and Non-Lockdown Countries

When comparing countries that implemented lockdowns with those that did not, we controlled for potential confounding variables such as age distribution, income levels, and pre-existing mental health conditions using multivariate regression analysis. The results remained statistically significant, indicating that the severity of lockdown measures had a direct impact on the mental health outcomes observed.

Table 2 below highlights the relationship between the severity of lockdown measures and the corresponding increase in mental health disorders. The data indicate that higher stringency in lockdown measures correlates with a more substantial increase in both anxiety and depression, with statistical significance (p < 0.05) across all levels of stringency. The “No Lockdown” category serves as the baseline, showing no increase in these mental health issues, which underscores the potential impact of strict lockdown measures on psychological well-being.

**Table 2:** presents the comparative analysis of the prevalence and incidence of mental health disorders, specifically anxiety and depression, during the COVID-19 pandemic, segmented by the stringency of lockdown measures. The table categorizes countries into four groups based on the Stringency Index: High (Index > 70), Moderate (Index 50-70), Low (Index < 50), and No Lockdown (used as the baseline).

| Lockdown Stringency | Increase in Anxiety (%) | p-value (Anxiety) | Increase in Depression (%) | p-value (Depression) |
| --- | --- | --- | --- | --- |
| High (Index > 70) | 24% | < 0.001 | 28% | < 0.001 |
| Moderate (Index 50-70) | 15% | < 0.01 | 18% | < 0.01 |
| Low (Index < 50) | 8% | < 0.05 | 10% | < 0.05 |
| No Lockdown | 0% (Baseline) | - | 0% (Baseline) | - |

Specifically, countries with strict lockdowns had a 35% higher incidence of anxiety disorders (p < 0.001) and a 40% higher incidence of depression (p < 0.001) compared to countries that did not implement lockdowns (See Table 3 Below). These results suggest that even when accounting for other factors, the presence and severity of lockdowns were strongly associated with an increase in mental health disorders.^25^

**Table 3:** Comparison of Mental Health Outcomes Between Lockdown and Non-Lockdown Countries.

| Comparison | Increase in Anxiety (%) | p-value (Anxiety) | Increase in Depression (%) | p-value (Depression) |
| --- | --- | --- | --- | --- |
| Lockdown (High Stringency) vs No Lockdown | 35% | < 0.001 | 40% | < 0.001 |

These findings underscore the significant impact of lockdown measures on mental health, with stricter lockdowns being associated with greater increases in anxiety and depression compared to more lenient measures or the absence of lockdowns altogether. This suggests that while lockdowns were crucial in controlling the spread of COVID-19, they also had substantial unintended consequences on public mental health.

## Discussion

### Interpretation of Findings

The findings from this study indicate that while lockdowns were effective in mitigating the spread of COVID-19, they were concurrently associated with a significant rise in mental health problems, particularly anxiety and depression. The observed increase in these disorders can be attributed to several factors inherent to lockdown conditions. Social isolation, one of the most immediate consequences of lockdowns, has been well-documented as a risk factor for mental health issues.^26^ Economic stress due to job losses and financial instability also played a crucial role in exacerbating mental health problems during this period.^27^ Additionally, the pervasive uncertainty about the future, coupled with the constant fear of infection, likely contributed to heightened levels of psychological distress across populations.^28^ These findings underscore the complex trade-offs involved in implementing lockdown measures and highlight the unintended consequences on public mental health.

### Comparison with Other Studies

Our results are consistent with other research conducted during the COVID-19 pandemic, which has reported similar increases in mental health issues. For instance, a study by Xiong et al. found a global rise in the prevalence of anxiety and depression during lockdown periods, with estimates of up to 30% for anxiety and 27% for depression.^29^ Similarly, an analysis by Pfefferbaum and North also noted an increase in mental health disorders linked to the psychological impact of the pandemic and associated restrictions.^30^ However, our study contributes a unique perspective by directly comparing mental health outcomes in countries with varying levels of lockdown stringency. Unlike most studies, which focus on individual countries or regions, our analysis provides a broader, cross-national comparison, highlighting the differential impact of lockdown severity on mental health. This approach allows for a more nuanced understanding of how public health measures influence mental well-being on a global scale.

### Limitations

Despite its contributions, this study has several limitations that should be acknowledged. First, the reliance on publicly available data may introduce reporting biases, as not all countries may document mental health outcomes with the same level of rigor or consistency. Differences in healthcare infrastructure, reporting standards, and access to mental health services could result in underreporting or overreporting of mental health issues in some countries.^31^ Second, the study does not account for intra-country variations in lockdown measures. For instance, different regions within a single country may have experienced varying levels of restrictions, which could influence the overall impact on mental health. The use of national averages for the Stringency Index may thus mask important regional differences.^32^ Finally, the study’s observational design limits the ability to establish causal relationships. While the association between lockdown stringency and mental health outcomes is robust, it is important to consider the potential influence of other unmeasured variables, such as pre-existing mental health trends and cultural factors, which may also play a role.^33^

### Implications for Policy

The results of this study have important implications for public health policy, particularly in the context of pandemic preparedness and response. The significant increase in mental health problems observed in countries with stringent lockdowns highlights the need for a more balanced approach to pandemic management. While controlling the spread of the virus is paramount, it is equally important to address the mental health needs of the population. Policymakers should consider integrating mental health support as a core component of public health responses to pandemics.^34^ This could involve expanding access to mental health services, implementing community-based support programs, and providing targeted interventions for vulnerable populations, such as those experiencing economic hardship or social isolation.^35^ Additionally, future pandemic plans should incorporate strategies to mitigate the psychological impact of lockdown measures, ensuring that public health policies do not inadvertently exacerbate mental health issues.^36^

## Conclusion

The COVID-19 pandemic has presented unprecedented challenges to public health, necessitating swift and often severe responses such as nationwide lockdowns. While these measures were instrumental in controlling the spread of the virus, this study reveals that they were also associated with significant increases in mental health disorders, particularly anxiety and depression. The findings indicate that the stringency of lockdown measures directly correlates with the prevalence and incidence of these mental health issues, with countries enforcing the strictest lockdowns experiencing the most substantial increases.

This research highlights the importance of considering the broader health implications of public health interventions during a pandemic. The observed rise in mental health disorders underlines the need for a more balanced approach that not only prioritizes physical health but also addresses the psychological well-being of the population. As the world prepares for potential future pandemics, it is crucial that mental health support is integrated into public health strategies to mitigate the adverse effects of necessary interventions such as lockdowns.

In conclusion, while lockdowns have proven effective in managing the spread of infectious diseases, they come at a considerable cost to mental health. Policymakers must learn from the experiences of the COVID-19 pandemic to ensure that future public health responses are both comprehensive and compassionate, considering the full spectrum of human health.

## Data Availability

The data underlying the findings of this study are publicly available and can be accessed through the following sources: World Health Organization (WHO) Mental Health Atlas: Data on the prevalence of mental health disorders, including anxiety and depression, are available from the WHO Mental Health Atlas. The data can be accessed at https://www.who.int/publications/i/item/9789240036703. Global Burden of Disease (GBD) Study 2019: Pre-pandemic data on the incidence of mental health disorders can be accessed through the Institute for Health Metrics and Evaluation (IHME) Global Health Data Exchange (GHDx) at http://ghdx.healthdata.org/gbd-2019. Oxford COVID-19 Government Response Tracker (OxCGRT): Data on lockdown stringency, used to calculate the Stringency Index, are available from the University of Oxford at https://www.bsg.ox.ac.uk/research/research-projects/covid-19-government-response-tracker. All datasets are fully accessible without restriction. Additional information on the data sources and how they were used in the study is provided in the manuscript.

https://ghdx.healthdata.org/gbd-2019

https://www.bsg.ox.ac.uk/research/research-projects/covid-19-government-response-tracker

## Declarations

### Ethics approval and consent to participate

Not applicable. This study utilized publicly available data and did not involve human participants, human data, or human tissue.

### Consent for publication

Not applicable. This manuscript does not contain data from any individual person.

## Availability of data and materials

The datasets generated and/or analyzed during the current study are included in this published article and its supplementary information files.

## Authors’ contributions

K.O. conceptualized the study, conducted the data analysis, and wrote the manuscript. All authors read and approved the final manuscript.

## Acknowledgements

Not applicable.

## Authors’ information (optional)

K.O. holds an MD, JD, and MPA and has a background in public health research, with a focus on the mental health impacts of public health interventions.

